# Plasma Amino Acids Metabolomics’ Profile Important for Glucose Management in Jordanian people with Type 2 Diabetes

**DOI:** 10.1101/2021.02.24.21252345

**Authors:** Abdelrahim Alqudah, Mohammed Wedyan, Esam Qnais, Hassan Jawarneh, Lana McClements

**Affiliations:** Department of clinical pharmacy and pharmacy practice, Faculty of pharmaceutical sciences, The Hashemite University, Zarqa, Jordan; Department of Biology and Biotechnology, Faculty of Science, The Hashemite University, Zarqa, Jordan; School of Life Sciences, Faculty of Science, University of Technology Sydney, Australia

**Keywords:** Type 2 diabetes, Amino acids, Jordanian

## Abstract

**Background:** The disturbances in plasma free amino acid metabolome in diabetes mellitus was studied before but not in Jordanian population. This study aimed to assess the association between type 2 diabetes (T2D) and amino acid metabolome in a representative group of people from Jordan.

**Methods:** Blood samples from 124 people with T2D and 67 age-, gender- and BMI-matched healthy controls were collected and assayed for glucose and HbA1c. Twenty one amino acids belonging to different categories (essential, non-essential, semi-essential, and metabolic indicators) were evaluated in both groups using amino acids analyser.

**Results:** Plasma free amino acids concentrations of total amino acids, total essential amino acids, total non-essential amino acids, and total semi-essential amino acids were not different in T2D compared to healthy controls. However, plasma concentrations of four essential amino acids (Leucine, Lysine, Phenylalanine, Tryptophan) were increased in the presence of T2D (Leucine, p<0.01, Lysine, p<0.001, Phenylalanine, p<0.01, Tryptophan, p<0.05). Conversely, amongst the non-essential amino acids, Alanine and Serine were reduced in type 2 diabetes (Alanine, p<0.01, Serine, p<0.001), whereas, Aspartate and Glutamate were increased in T2D compared to healthy control plasma (Aspartate, p<0.001, Glutamate, p<0.01). A semi-essential amino acid, Cystine, was also increased in T2D compared to healthy controls (p<0.01). Citrulline, a metabolic indicator amino acid, demonstrated lower plasma concentration in T2Dcompared to healthy controls (p<0.01).

**Conclusion:** Several amino acids from different categories are dysregulated in T2D, which could be used as a therapeutic target to improve T2D management and its complications.

## Introduction

According to the World Health Organisation (WHO) the number of people with diabetes increased from 108 million in 1980 to 422 million in 2014, and the prevalence of diabetes has been rising rapidly in the middle- and low-income countries (*Diabetes*, 2020). Approximately 1.6 million deaths in 2016 were directly caused by diabetes, with half of all deaths occurring before the age of 70 years (2). In 2004, the prevalence of diabetes among Jordanian population aged between 25 and 70 was 17.1% which was increased to 23.7% in 2017 which was higher than the world average. This increase in the prevalence of diabetes in Jordan might be due to poor diet and technological advances that have changed the lifestyle to a more sedentary pattern (3). Diabetes is a metabolic disorder characterised by chronic hyperglycaemia, affecting carbohydrate, fat, and protein metabolism leading to abnormal insulin secretion, insulin resistance, or both (4). Diabetes mellitus is characterised by the following symptoms: thirst, polyuria, blurred vision, and weight loss. These symptoms can be quiescent for a prolonged period of time resulting in retinopathy, nephropathy, neuropathy and cardiovascular complications due to persistent hyperglycaemia, occurring before the diagnosis of diabetes mellitus is made (5). The most frequent form of diabetes is type 2 diabetes (T2D). T2D is characterised by hyperglycaemia, insulin resistance and relative insulin deficiency, often at later stages in life (6). The causes of T2D are still not fully understood; however, high body mass index (BMI), age, ethnicity and family history have all been linked to the epidemiology of T2D (7). Other factors have also shown some association with diabetes including poor diet and nutrition, physical inactivity, impaired glucose tolerance, smoking, and past history of exposure of the unborn child to high glucose during pregnancy (maternal gestational diabetes) (8). Furthermore, recent evidence has suggested that high sugar intake is associated with risk of T2D (9).

As amino acids (AA) are the building units of proteins with critical roles in gene expression, cell signalling, reproduction, neurotransmission, metabolism, oxidative stress, pain control, inflammatory responses, and detoxification (10). Human cells can synthesize several amino acids including alanine (Ala), asparagine (Asn), aspartate (Asp), glutamate (Glu), glutamine (Gln), glycine (Gly), proline (Pro) and serine (Ser) that are referred to as non-essential amino acids. On the other hand, some amino acids are obtained from the diet and referred to as essential amino acids. These include histidine (His), isoleucine (ILe), Leucine (Leu), lysine (Lys), methionine (Met), phenylalanine (Phe), threonine (Thr), tryptophan (Trp), and valine (Val). Semi-essential amino acids including arginine, cysteine (Cys), and tyrosine (Tyr) can be synthesised endogenously, however in insufficient amounts, therefore, these are required partially to be obtained from the diet (11). Other amino acids with the metabolism conversion capacity or dietary adequacy have a role as metabolic indicators of impaired metabolism and these include ornithine (Orn) and citrulline (Cit) (12). Metabolomics technologies have found a strong association between some amino acids and insulin resistance (13). Furthermore, a number of published studies reported that certain pathological conditions including metabolic and cardiovascular diseases are associated with disturbances in plasma free amino acids (PFAA) metabolism (14), hence these amino acids could be utilised as potential target for therapeutic intervention in such conditions including diabetes. Although, aberrant PFAA concentrations have been reported previously in in people with T2D in different countries, no comprehensive study was conducted to assess the PFAA concentrations in T2D in Jordanian cohorts (12,15). Therefore, this study aims to determine amino acid metabolome in people with T2D compared to healthy controls in Jordan hence providing a better understanding of the role of individual amino acids in this metabolic disorder that could improve the management and reduce diabetic complications.

## Methods

### Study population

Participants with T2Dor healthy controls referred to Endocrinology Department at the Princess Bassma public hospital and King Abdullah University hospital were consented from January to April 2019 before donating a blood sample. A total of 124 participants with T2D and 67 healthy controls were recruited in this study. Inclusion criteria was set as follows: T2D diagnosed at least 6 months prior to sample collection, residing in Jordan, age >30 years old, and treatment regimen that included metformin only. Exclusion criteria included the following conditions: type 1 diabetes and chronic complications of T2D (nephropathy, retinopathy, and cardiovascular disease).

### Ethical consideration

Ethical approval for this study was obtained from the Institutional Review Board (IRB number 6/7/2017/2018) at the Hashemite University, Zarqa, Jordan, and all procedures followed were in accordance with the 1964 Declaration of Helsinki.

### Clinical and laboratory assessments

BMI was calculated using self-reported weight (kg) divided by height in meters squared (kg/m2). Blood pressure was measured by trained nurses 3 times using the right arm after a 10-minutes rest period. Participants were asked to fast for 8 hours prior to blood sample collection. Venous blood samples were collected from peripheral vein in heparinized collection tubes. Blood samples were mixed gently, centrifuged at 3,0000 rpm at 4°C for 15 minutes and the plasma layers were kept frozen at −80°C for further analysis. HbA1c values were measured using commercially available kit (Wondfo, China) and blood glucose levels were assessed using colorimetric detection kit (Biolabo, France) according to the manufacturer’s instructions.

### Amino acid analysis

To reduce any bias introduced prior to analysis, samples were given a unique code and were blinded. Plasma samples were prepared and deproteinized using amino acid sample preparation kit (MembraPure Gmbh, Germany) according to the manufacturer’s instructions. Total amino acid profiling was carried out using Amino Acid Analyzer (ARACUS, membraPure Gmbh, Germany).

### Statistical analysis

Data were represented as mean ± SD. All analysed parameters were tested for normality of the data using the Kolmogorov-Smirnov test. Unpaired student t-test was used to test the significance of data using Prism 5 software (Graphpad software, USA). P<0.05 was taken as the cut off value for significance.

## Results

### Baseline characteristics

Plasma samples were collected from 124 patients with type 2 diabetes, and 64 samples from healthy controls. Baseline characteristics are shown in **Table 1**. No statistically significant differences in age, gender, BMI, systolic and diastolic blood pressure were noted between the study group and their controls. As expected, patients with T2D had higher blood concentration of fasting blood sugar (FBS) and HbA1c compared to healthy controls (**Table 1**, p<0.001).

**Table 1:**
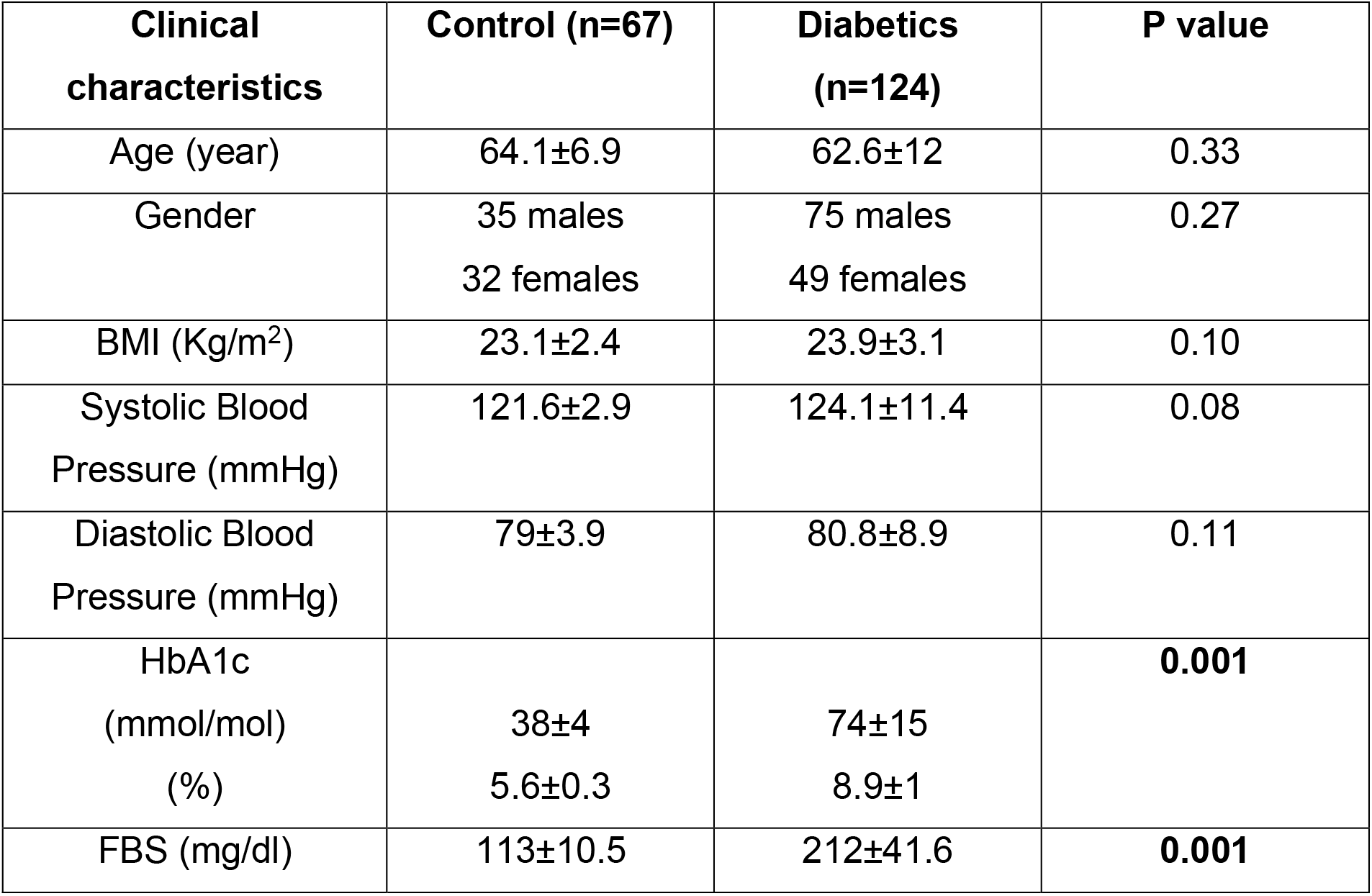
Clinical characteristics for the participants

| **Clinical characteristics** | **Control (n=67)** | **Diabetics (n=124)** | **P value** |
| --- | --- | --- | --- |
| Age (year) | 64.1±6.9 | 62.6±12 | 0.33 |
| Gender | 35 males<br>32 females | 75 males<br>49 females | 0.27 |
| BMI (Kg/m<sup>2</sup>) | 23.1±2.4 | 23.9±3.1 | 0.10 |
| Systolic Blood Pressure (mmHg) | 121.6±2.9 | 124.1±11.4 | 0.08 |
| Diastolic Blood Pressure (mmHg) | 79±3.9 | 80.8±8.9 | 0.11 |
| HbA1c (mmol/mol) | 38±4 | 74±15 | **0.001** |
| (%) | 5.6±0.3 | 8.9±1 |  |
| FBS (mg/dl) | 113±10.5 | 212±41.6 | **0.001** |

### Plasma levels of the major categories of amino acids in Jordanian patients with T2D

The total PFAA concentrations were assessed in order to determine any disturbances in amino acid metabolism in this group of Jordanian participants as a result of T2D. No statistically significant difference was observed in the total amino acids concentrations between T2D participants and healthy controls (**Fig. 1A**). Also, total essential, non-essential and semi-essential amino acids PFAA concentrations were not statistically different between these two groups (**Fig. 1B, C&D, respectively**).

**Figure 1.**
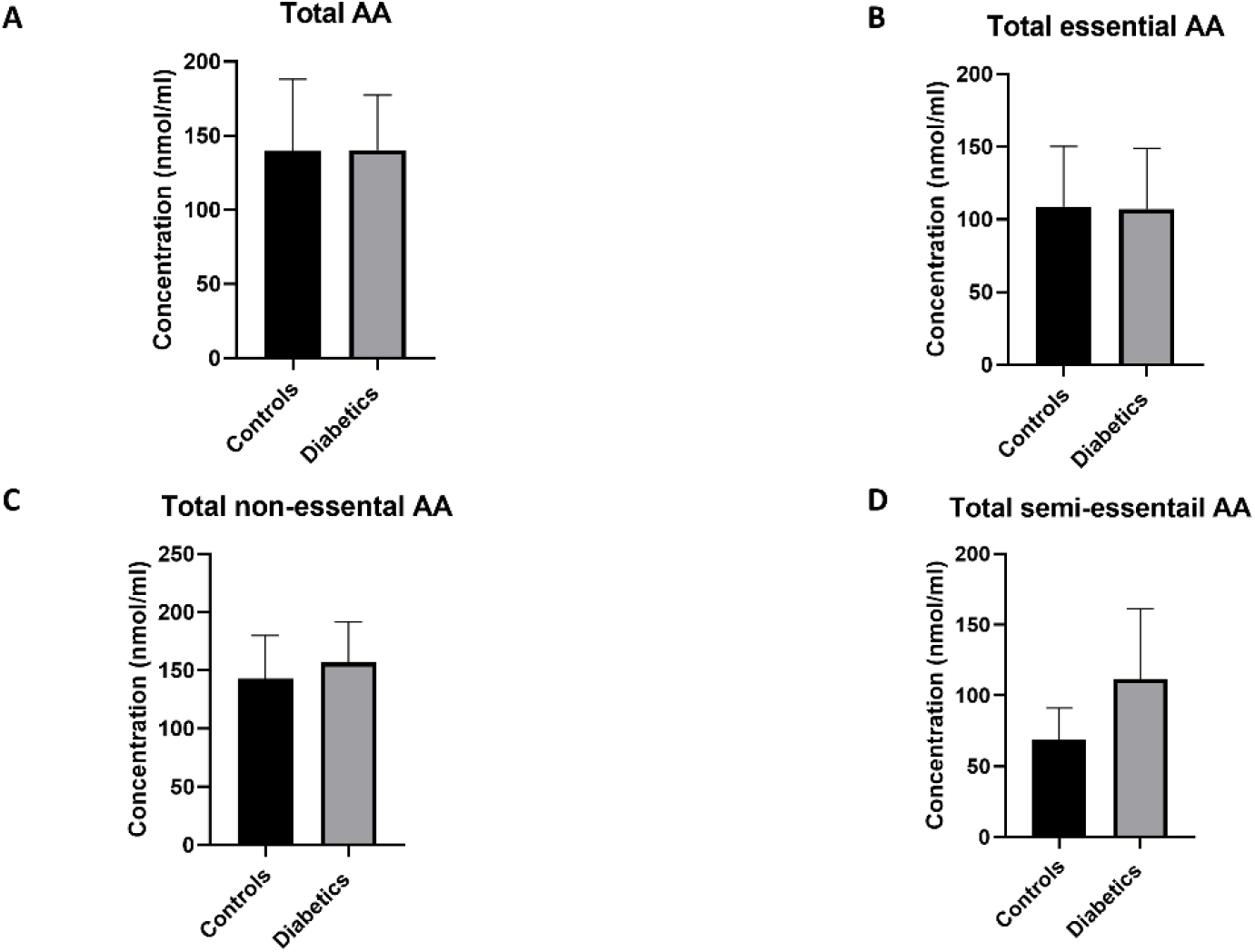
Plasma concentration of total amino acids in type 2 diabetes (T2D). No significant difference in free plasma concentration of total amino acids (**A**), essential amino acids (**B**), non-essential amino acids (**C**), and semi-essential amino acids (**D**) between patients with T2D and healthy controls. Plasma samples from T2D patients (n=124) or healthy controls (n=67) were prepared and deproteinized before total amino acid profiling was carried out (unpaired independent t-test).

### Effect of T2D on essential amino acids

By analysing individual essential PFAA concentration, no significant difference in free plasma concentrations of His, ILe, Met, Thr, and val was found between patients with T2D and controls (**Fig. 2**). Furthermore, no significant correlation between plasma free concentrations of these amino acids and fasting blood sugar (FBS) and HbA1c levels in T2D (FBS, **Table 2;** HbA1c, **Table 3**). However, significant increase in free plasma concentrations of Leu, Lys, Phe and Trp was noted in patients with T2D compared to their healthy controls (**Fig. 2**; Leu, Lys, Phe, p<0.01, Trp, p<0.05). These results were consistent when correlation was performed between free plasma concentrations of essential amino acids and FBS. Leu, Lys, Phe, and Trp free plasma concentrations were positively correlated with FBS (**Table 2**; Leu, r=0.53, p=0.004; Lys, r=0.395, p=0.03; Phe, r=0.42, p=0.006; Trp, r=0.675, p=0.0001). Similarly, Leu, Lys, Phe, and Trp free plasma concentrations were positively correlated with HbA1c (**Table 3**; Leu, r=0.44, p=0.02; Lys, r=0.43, p=0.012; Phe, r=0.49, p=0.001; Trp, r=0.59, p=0.001).

**Figure 2.**
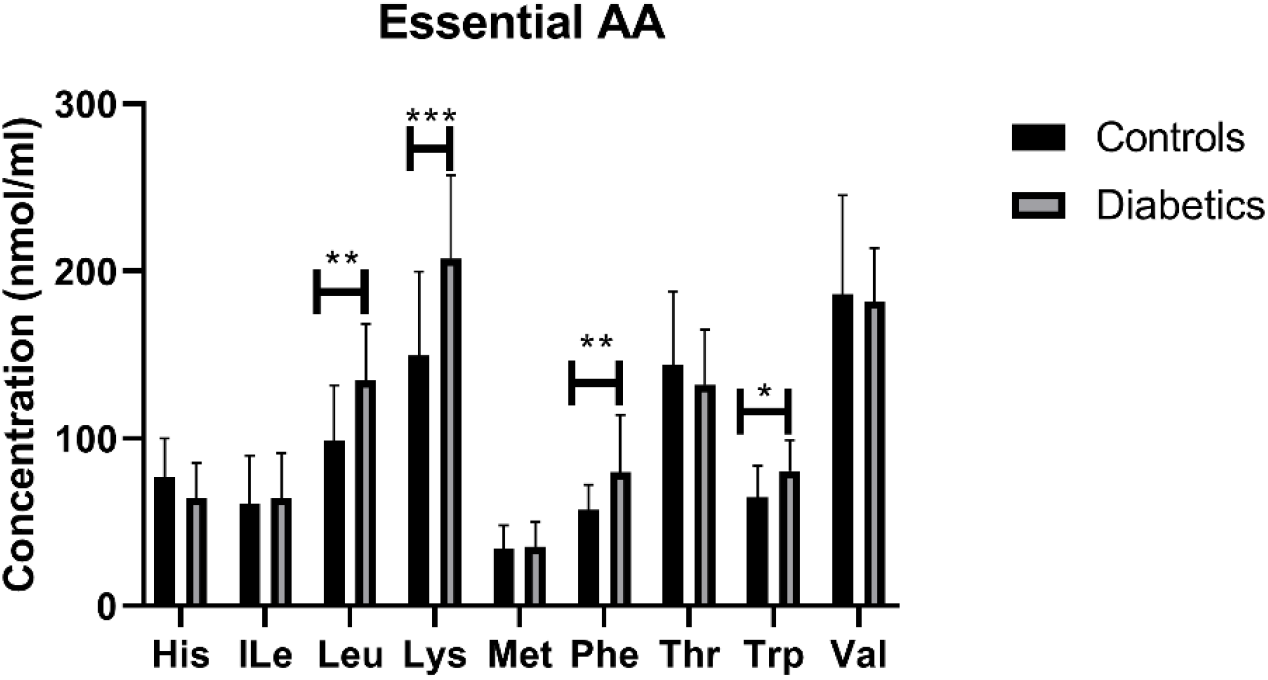
Plasma concentration of essential amino acids in type 2 diabetes (T2D). Statistically significant increase in Leu, Lys, Phe, and Trp was found between patients with T2D and healthy controls. Plasma samples from T2D participants (n=124) or healthy controls (n=67) were prepared and deproteinized before total amino acid profiling was carried out (unpaired independent t-test, *<0.05, **<0.01).

**Table 2:** Correlation analysis between FBS and amino acids concentrations in type 2 diabetes

| Amino acid | Correlation (r) | P value |
| --- | --- | --- |
| His | 0.050 | 0.80 |
| ILe | 0.164 | 0.41 |
| Leu | 0.53 | **0.004** |
| Lys | 0.395 | **0.03** |
| Met | -0.161 | 0.42 |
| Phe | 0.42 | **0.006** |
| Thr | -0.163 | 0.18 |
| Trp | 0.675 | **0.0001** |
| Val | -0.158 | 0.58 |
| Ala | -0.46 | **0.01** |
| Asn | -0.019 | 0.88 |
| Asp | 0.886 | **0.001** |
| Glu | 0.550 | **0.001** |
| Gln | -0.041 | 0.87 |
| Gly | -0.178 | 0.24 |
| Ser | -0.555 | **0.0004** |
| Arg | -0.365 | 0.09 |
| Cys | 0.569 | **0.005** |
| Tyr | -0.261 | 0.14 |
| Orn | 0.312 | 0.08 |
| Cit | -0.438 | **0.01** |

**Table 3:**
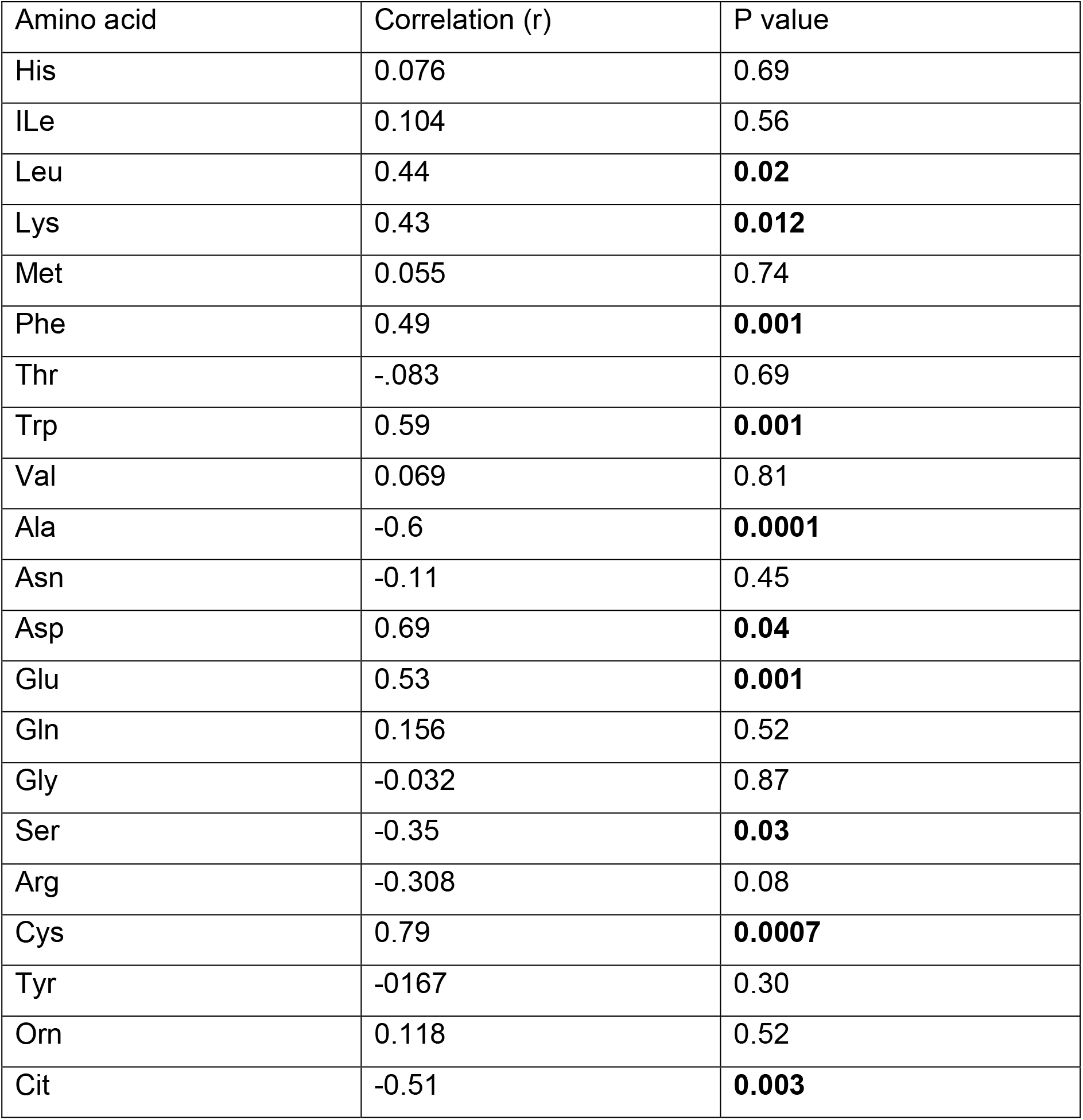
Correlation analysis between HbA1c and amino acids concentrations in T2D

| Amino acid | Correlation (r) | P value |
| --- | --- | --- |
| His | 0.076 | 0.69 |
| ILe | 0.104 | 0.56 |
| Leu | 0.44 | **0.02** |
| Lys | 0.43 | **0.012** |
| Met | 0.055 | 0.74 |
| Phe | 0.49 | **0.001** |
| Thr | -.083 | 0.69 |
| Trp | 0.59 | **0.001** |
| Val | 0.069 | 0.81 |
| Ala | -0.6 | **0.0001** |
| Asn | -0.11 | 0.45 |
| Asp | 0.69 | **0.04** |
| Glu | 0.53 | **0.001** |
| Gln | 0.156 | 0.52 |
| Gly | -0.032 | 0.87 |
| Ser | -0.35 | **0.03** |
| Arg | -0.308 | 0.08 |
| Cys | 0.79 | **0.0007** |
| Tyr | -0.167 | 0.30 |
| Orn | 0.118 | 0.52 |
| Cit | -0.51 | **0.003** |

### Effect of T2D on non-essential amino acids

In terms of the non-essential amino acids, no statistically significant difference was noted in free plasma concentrations of Asn, Gln, and Gly between patients with T2D compared to controls **(Fig. 3**); the correlation between free plasma concentrations of these amino acids and FBS and HbA1c levels in T2D was also not statistically significant (**Table 2 and Table 3, respectively**). Plasma levels of Ala and Ser were significantly decreased in patients with T2D compared to healthy controls (**Fig. 3**, Ala, p<0.01, Ser, p<0.001), whereas, plasma levels of Asp and Glu showed a significant increase in patients with T2D compared to healthy controls (**Fig. 3**; Asp, p<0.001, Glu, p<0.01). Similarly, Ala and Ser plasma concentrations in patients with T2D were negatively correlated with FBS (**Table 2**; Ala, r=-0.46, p=0.01; Ser, r=-0.555, p=0.0004). In addition, HbA1c levels were negatively correlated with Ala and Ser levels (**Table 3**; Ala, r=-0.6, p=0.0001; Ser, r=-0.35, p=0.03), whilst Asp and Glu concentrations were positively correlated with FBS (**Table 2**; Asp, r=0.886, p=0.001; Glu, r=0.55, p=0.001) and HbA1c levels (**Table 3**; Asp, r=0.69, p=0.04; Glu, r=0.53, p=0.001) in T2D.

**Figure 3.**
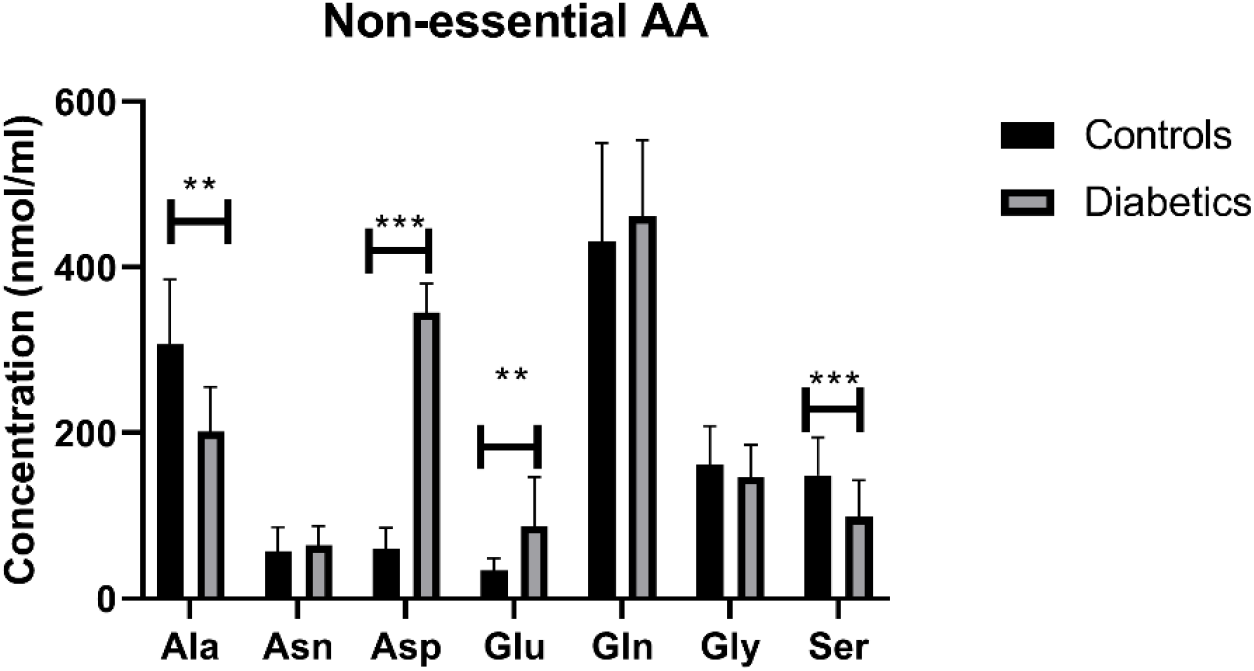
Plasma concentration of non-essential amino acids in type 2 diabetes (T2D). Ala concentration was reduced in T2D, whereas Asp, Glu, and Ser concentrations were increased compared to healthy controls. Plasma samples from T2D participants (n=124) or healthy controls (n=67) were prepared and deproteinized before total amino acid profiling was carried out (unpaired independent t-test, **<0.01, ***<0.001).

### Effect of T2D on semi-essential and metabolic indicators amino acids

There was no statistically significant difference in free plasma concentrations of semi-essential amino acids, Arg and Tyr between T2D and healthy control plasma samples (**Fig. 4A**). In addition, the correlation between free plasma concentrations of these amino acids and FBS (**Table 2**) and HbA1c (**Table 3**) levels in T2D was not demonstrated (**Table 2**). However, free plasma concentration of Cys showed a significant increase in patients with T2D compared to healthy controls (**Fig. 4A**, p<0.01). Moreover, Cys free plasma concentration was positively correlated with FBS (**Table 2**; r=0.569, p=0.005) HbA1c (**Table 3**; r=0.79, p=0.0007) levels in T2D.

**Figure 4.**
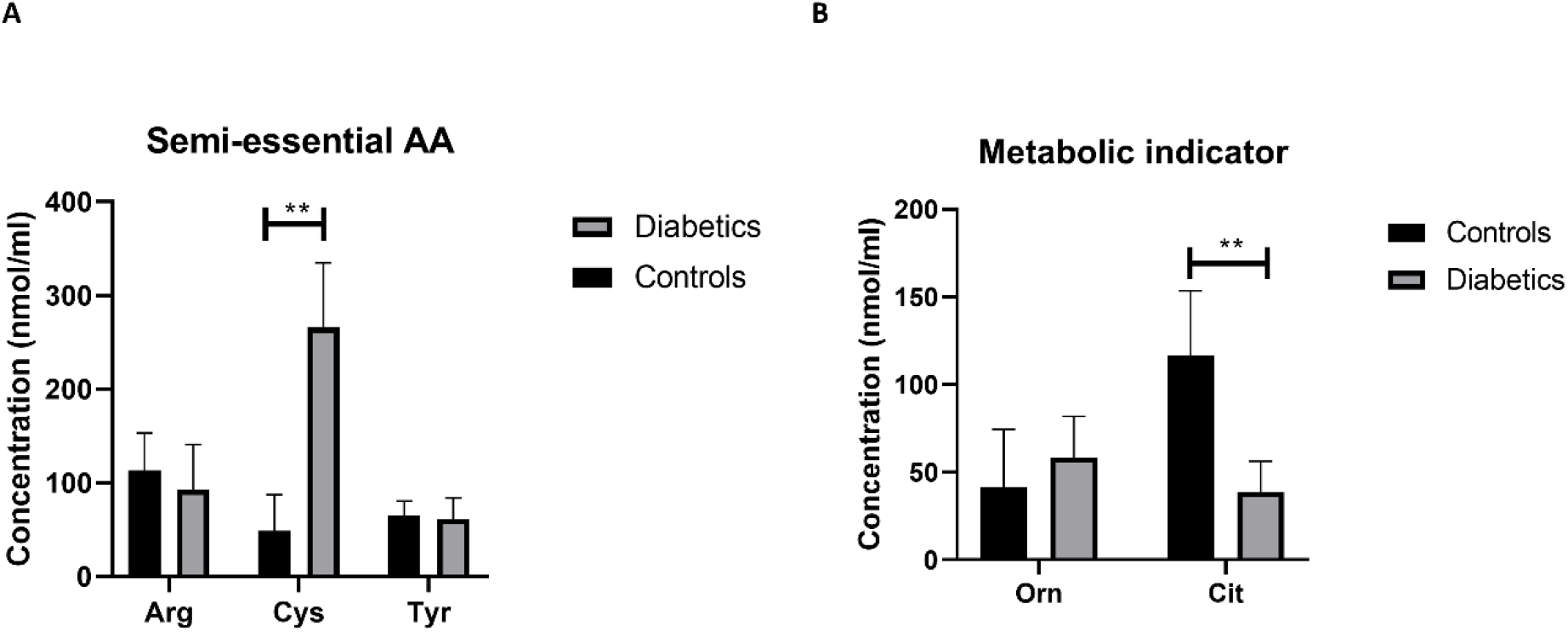
Plasma concentration of semi-essential (A) and metabolic indicators (B) amino acids in type 2 diabetes (T2D). Cys plasma concentration was increased in T2D participants compared to healthy controls (**A**). Cit plasma concentration was reduced in T2D compared to healthy control (**B**). Plasma samples from T2D participants (n=124) or healthy controls (n=67) were prepared and deproteinized before total amino acid profiling was carried out (unpaired independent t-test, **<0.01).

With regards to metabolic indicators amino acids, Orn free plasma concentration did not show a significant difference between patients with T2D and controls (**Fig. 4B**), and the correlation between Orn free plasma concentration and FBS (**Table 2**) and HbA1c (**Table 3**) levels in T2D was not statistically significant. Interestingly, Cit free plasma concentration demonstrated a significant decrease in patients with T2D compared to healthy controls (**Fig. 4B**, p<001). In addition, Cit free plasma concentration was negatively correlated with FBS (Table 2; r=-0.438, p=0.01) and HbA1c (**Table 3**; r=-0.51, p=0.003) levels in T2D.

## Discussion

To our knowledge, this is the first study describing the association between T2D and plasma free amino acids concentrations in Jordanian participants that sheds further light on the role of specific amino acid metabolism in T2D and glucose management. In this study, we conducted introductory metabolomic analysis of 124 patients with T2D and 67 healthy control individuals. A recent study conducted in China reported that there was a significant increase in plasma concentration of six essential amino acids (ILe, Leu, Lys, Phe, Trp and Val), three non-essential and semi-essential amino acids (Ala, Glu and Tyre) in T2D, and this increase was associated with higher risk of T2D prevalence and/or incidence (16). Aligned to the findings of this study, here we demonstrate that Leu, Lys, Phe, Trp, and Glu plasma concentrations were significantly increased in patients with T2D compared to controls, and positively associated with poor glucose management. Furthermore, some reports suggested that essential amino acids including branched chain amino acids (BCAA) ILe, Leu, and Val, obtained from the diet and metabolised in adipose tissues and skeletal muscles, are strongly associated with insulin resistance (17). Our results demonstrated that Leu plasma concentration was significantly increased in patients with T2D reflective of worse glucose management. Low levels of Leu were reported to improve insulin sensitivity in the liver by activating general control non-derepressible (GCN)2 and decreasing the activity of rapamycin/S6K1 signalling, in addition to AMPK activation, suggesting that high levels of Leu is strongly associated with insulin resistance (18). Additionally, Phe, an aromatic amino acid (AAA), was increased in participants with T2D in our study, which is consistent with several previous studies showing that Phe concentration was elevated in response to obesity and insulin resistance (19). Moreover, our results showed that plasma concentration of another AAA, Trp, was also increased in T2D. Trp is implicated in tryptophan-kynurenine and tryptophan-methoxyindole metabolic pathways involved in the production of a number of active metabolites including kynurenine, kynurenic acid and serotonin. Any disturbances in this metabolic pathways are likely associated with T2D pathophysiological mechanisms (20). Furthermore, Lys, an essential amino acid, was increased in patients with T2D in this study. Razquin et al, demonstrated that Lys levels is associated with higher risk of T2D. In addition, subjects with T2D and high levels of Lys appear to have increased risk of cardiovascular disease (21)

On the other hand, Ala, a non-essential amino acid, was reported to be increased in hyperinsulinemia conditions in diabetes, and supplementation of Ala improved glucagon response to hypoglycaemia events in diabetes (22). In this study, Ala plasma concentration was significantly reduced in T2D patients which could indicate that the patients in this study might be at higher risk of hypoglycaemic events. Furthermore, our results showed that plasma concentrations of Asp and Glu, acidic amino acids, were increased in patients with T2D, which is in line with previous reports showing that acidic amino acids plasma concentration was higher in patients with T2D compared to controls (23). Recent study reported that adiponectin concentration is negatively correlated with Asp and Glu concentrations with a potential to increase the risk of cardiovascular disease (24). Additionally, our results indicate that Ser, a non-essential amino acid, plasma concentration was significantly decreased in patients with type 2 diabetes. These results were consistent with a Japanese’s study reporting that a cohort who developed diabetes within four years of follow up had a lower Ser concentration which was significantly related to the development of metabolic syndrome (25).

In our study, Cys, a semi-essential amino acid, was elevated in T2D compared to controls and associated with high HbA1c. Previous reports show that Cys plasma concentration was positively correlated with obesity and insulin resistance, and that higher levels of Cys could lead to obesity (26). Moreover, higher concentration of Cys was positively correlated with markers of inflammation including C-reactive protein (CRP) and tumor necrosis factor-α (TNF-α), suggesting higher risk of diabetes complications (27). Cit is a key modulator of urea cycle in the liver and kidneys that can be synthesized from Orn by the intestines (28). In addition, Cit can contribute to Arg synthesis before it is converted to nitric oxide (NO) by endothelial nitric oxide synthase (eNOS) (29). NO has an essential role in maintaining endothelial function and vasodilation, in addition to regulating insulin sensitivity (30). Our results indicate a significant reduction in Cit free plasma concentration that could suggest that there is a reduction in NO and increased risk of future cardiovascular disease in these participants. Restoring Cit levels through supplementation could potentially prevent or delay cardiovascular complications in people with T2D.

Limitations of this study include the following aspects of the study: (i) the levels of amino acids are partially influenced by the dietary habits, the information we did not obtain, and (ii) the sample size that was small, cross-sectional and from Jordanian population only, hence limiting the application of our results. Nevertheless, our results aligned with the published literature warranting further investigations into abovementioned amino acid with impaired metabolism using larger longitudinal cohorts of people with T2D and healthy controls.

In conclusion, this study demonstrated that specific amino acids with different important biological roles are dysregulated in T2D impacting on glucose management and with a potential to increase the risk of future diabetic complications, which could be used as a therapeutic target to improve T2D management and its complications.

## Data Availability

The datasets generated during and/or analysed during the current study are available from the corresponding author on reasonable request.

## Acknowledgment

The authors would like to thank all the participants in this study for their cooperation and the staff of Princess Bassma governmental hospital and King Abdullah University hospital for their technical assistance.

